# Prevalence, Hallmark-Based Mechanisms, and Risk Factors of Peripheral Diabetic Neuropathy: A Systematic Review and Meta-Analysis

**DOI:** 10.64898/2026.08.02.26359538

**Authors:** Dipamoy Datta, Dwaipayan Saha, Riddhiman Ghosh, Arjun Baidya, Bhaswati Ganguli, Subhra Prakash Hui

## Abstract

Diabetic peripheral neuropathy (DPN) imposes a substantial global burden, yet its mechanistic underpinnings and integrated risk architecture remain incompletely characterized. This systematic review and meta-analysis of 74 studies across 24 countries (83,560 participants) provides the most comprehensive quantitative synthesis of DPN prevalence, hallmark-based pathogenesis, and multi-covariate risk profiling to date. The global pooled DPN prevalence was 57.42% (95% CI: 48.56–66.05), with continent-specific gradients: Americas (74.52%), Europe (59.95%), and Asia (45.44%). Hallmark-stratified subgroup analyses encompassing neuronal damage, metabolic dysregulation, neuroinflammation, and microvascular alteration identified microvascular alteration as the sole statistically significant mechanistic determinant (Q = 15.78, p = 0.0004), with prevalence escalating monotonically from 53% to 92% across increasing hallmark severity scores, constituting a compelling dose-response relationship. Risk factor meta-analysis of 27 covariates identified 12 significant determinants, including the novel meta-analytic confirmation of peripheral vascular disease (OR: 3.70; highest effect size), male sex (OR: 1.52), and height (OR: 1.26) as independent DPN risk factors. Notably, HbA1c, BMI, and blood pressure were non-significant, challenging glycemia-centric paradigms. These findings collectively support a precision medicine framework grounded in hallmark-stratified phenotyping and mechanism-targeted pharmacotherapy for DPN.

## INTRODUCTION

Diabetic peripheral neuropathy (DPN) is the most prevalent microvascular complication of diabetes mellitus, affecting approximately 50% of the global diabetic population [1–3]. DPN presents as a length-dependent, progressive peripheral nervous system (PNS) disorder, manifesting as a debilitating spectrum of sequelae: neuropathic pain, sensory deficit, motor dysfunction, postural instability, heightened fall risk, fragility fractures, diabetic foot ulceration (DFU), and premature mortality [1–5]. DPN’s role in precipitating DFU is particularly consequential, substantially amplifying risk for limb infection, structural foot deformity, micro-traumatism, and lower-extremity amputation [2,5,6]. Approximately 25% of DPN patients develop a painful phenotype — painful DPN (PDPN) — characterized by chronic neuropathic pain of variable severity, imposing additional psychosocial burden including depression, anxiety, and sleep disturbance [2–5,16].

Mechanistically, DPN is best characterized as a neurodegenerative disease of the PNS, primarily targeting sensory axons in a distal-to-proximal, “dying-back” pattern [3,7]. Despite decades of investigation, the complete pathophysiological basis of DPN remains incompletely elucidated, critically impeding the development of mechanism-targeted, curative pharmacotherapies and perpetuating the chronic underassessment and under treatment of sensory neuronal loss and neuropathic pain [2,3,5,7–9]. Current symptomatic therapies provide only partial pain relief, fail to reverse established structural nerve injury, and carry appreciable long-term adverse effect burdens [2–5,7,8]. The clinical phenotype of DPN exhibits marked inter-patient heterogeneity in symptom constellation, pain quality, and disease trajectory, rendering universal therapeutic strategies fundamentally inadequate [3,10].

Confronting these challenges demands individualized, phenotype-stratified, mechanism-based therapeutic approaches the precision medicine paradigm for DPN [3,5,7,10]. A prerequisite for this approach is a comprehensive mechanistic framework integrating the dominant pathophysiological hallmarks driving DPN initiation and progression. The failure of intensive glycemic control alone to prevent DPN including in well-controlled patients with HbA1c < 5.4% cogently underscores the involvement of pathophysiological mechanisms beyond hyperglycemia and reinforces the imperative for a multi-hallmark mechanistic model [3,5,17]. Multiple risk factors including patient age, diabetes duration, HbA1c, fasting plasma glucose (FPG), body mass index (BMI), diastolic blood pressure (DBP), diabetic retinopathy (DR), and tobacco use modulate DPN susceptibility and complication severity [3,5,6,13,16].

Critically, neither an established quantitative computational framework for DPN pathogenesis nor mechanistic meta-analytic evidence from clinical data currently exist [11–15]. Addressing this knowledge gap, we previously proposed a four-hallmark mechanistic framework of DPN pathogenesis encompassing neuronal damage, microvascular alteration, metabolic dysregulation, and neuroinflammation under the auspices of the Indian Diabetes Precision Medicine Initiative [18]. Building upon this theoretical construct, the present study constitutes the first systematic review and meta-analysis to quantitatively validate this hallmark-based mechanistic model using DPN clinical data. The specific objectives are: (i) to estimate the global pooled prevalence of DPN using random-effects models; (ii) to determine continent-specific DPN prevalence through subgroup meta-analysis; (iii) to assess whether study-level hallmark severity scores explain inter-study prevalence heterogeneity; (iv) to systematically evaluate clinical and demographic risk factors for DPN; and (v) to synthesize pooled odds ratios for established and novel DPN risk covariates.

## METHODS

### Search Strategy

A systematic electronic search was conducted across five databases: PubMed, Google Scholar, Medline, Cochrane Library, and CINHAL, following Cochrane Collaboration recommendations [19]. Searches were initiated in June 2023 and extended through November 2024. The search strategy was structured around the four-hallmark pathophysiological framework [18], integrating MeSH terms with free-text keywords via Boolean operators (AND/OR). Key search terms encompassed: diabetic peripheral neuropathy, neuronal damage, metabolic dysregulation, microvascular alteration, neuroinflammation, hyperglycemia, dyslipidemia, axonal loss, segmental demyelination, nerve fiber density, sensory neuronal degeneration, neurotransmitter imbalance, endothelial dysfunction, capillary basement membrane widening, and pericyte loss.

### Study Selection

Studies were eligible if they: (a) reported mechanistic evidence relevant to DPN pathogenesis; (b) clinically confirmed diabetes mellitus and DPN; (c) utilized cross-sectional, case-control, or cohort designs; (d) were hospital-, population-, or community-based; (e) were original full-text peer-reviewed articles published in English between 1989 and 2024; and (f) included patients of any age. Exclusion criteria encompassed: incomplete primary pathogenesis data; reviews, commentaries, editorials, or conference abstracts; animal or in vitro experimental models; non-diabetic etiological neuropathies (e.g., chronic inflammatory demyelinating polyneuropathy, hereditary neuropathy, alcoholic polyneuropathy, paraneoplastic neuropathy, or collagen vascular disease-associated neuropathy); and absence of mechanistic insights pertinent to DPN.

### Data Extraction

Standardized data extraction was performed into Microsoft Excel. Extracted variables included: first author, country, publication year, study design, sample size, DPN patient count, neuropathy type, diabetes type, diabetes duration, patient age, diagnostic criteria, DPN grouping criteria, hallmark involvement categories, associated pathophysiological features, HbA1c (%), FPG (mg/dL), BMI (kg/m²), systolic blood pressure (SBP), DBP, total cholesterol, HDL-cholesterol, LDL-cholesterol, triglycerides (mg/dL), and reported odds ratios (ORs) with corresponding 95% confidence intervals (CIs).

### Methodological Quality Assessment and Risk of Bias

Methodological quality and risk of bias for all included observational studies were evaluated using the Newcastle–Ottawa Scale (NOS) [20,21], assessing three domains: participant selection, comparability of study groups, and outcome ascertainment with follow-up adequacy. NOS scores range from 0 (poorest quality) to 9 (highest quality), with scores of 7–9 indicating low risk of bias (“good” quality), 4–6 indicating moderate risk (“fair” quality), and 0–3 indicating high risk of bias (“poor” quality).

### Statistical Analysis

Prevalence data were synthesized using random-effects meta-analysis. To stabilize variance prior to pooling, the Freeman–Tukey double arcsine transformation was applied. Between-study variance (τ²) was estimated via restricted maximum likelihood (REML). Heterogeneity was quantified using τ² and I². Given the consistently high heterogeneity observed, random-effects estimates were considered primary. For risk factor covariates reported across multiple studies, odds ratios were log-transformed prior to meta-analysis. Pooled ORs and corresponding 95% CIs were computed under a normal–normal hierarchical random-effects model with REML estimation of τ², and CIs were constructed using the Hartung–Knapp adjustment. All analyses were conducted in R (version 4.x) using the meta and metafor packages.

### Subgroup Analysis

A priori subgroup analyses were conducted to explore sources of heterogeneity in DPN prevalence across study-level hallmark severity indicators, with subgroup thresholds defined to maintain a minimum of five contributing studies per subgroup. Subgroup definitions were Neuronal damage: 0, 1, ≥2 features; Microvascular alteration: 0, 1, ≥2 features; Metabolic dysregulation: 0, 1, 2 features; Neuroinflammation: 0, 1 feature. Within each subgroup, pooled prevalence was estimated using random-effects models. Tests for subgroup differences were performed under both common-effect and random-effects assumptions, with primary interpretation based on random-effects results.

## RESULTS

### Study Selection and Characteristics

The systematic search identified 2,032 potentially relevant articles from five electronic databases (PubMed, n=1,179; Google Scholar, n=130; Medline, n=68; Cochrane, n=36; CINHAL, n=12) and manual citation screening (n=607). Following removal of 794 duplicates, 1,238 articles underwent title and abstract screening, with 533 excluded for misalignment with the pre-defined DPN pathophysiological framework [18]. Full-text review of the remaining 705 articles led to further exclusion of 390 records (review articles/editorials, n=38; animal studies, n=61; incomplete datasets, n=79; irrelevant clinical outcomes, n=101; insufficient diagnostic criteria, n=39; non-diabetic neuropathies, n=72). A final cohort of 74 studies met all inclusion criteria and was retained for meta-analysis (Fig. 1) (Supplementary Table S1).

**Figure 1.**
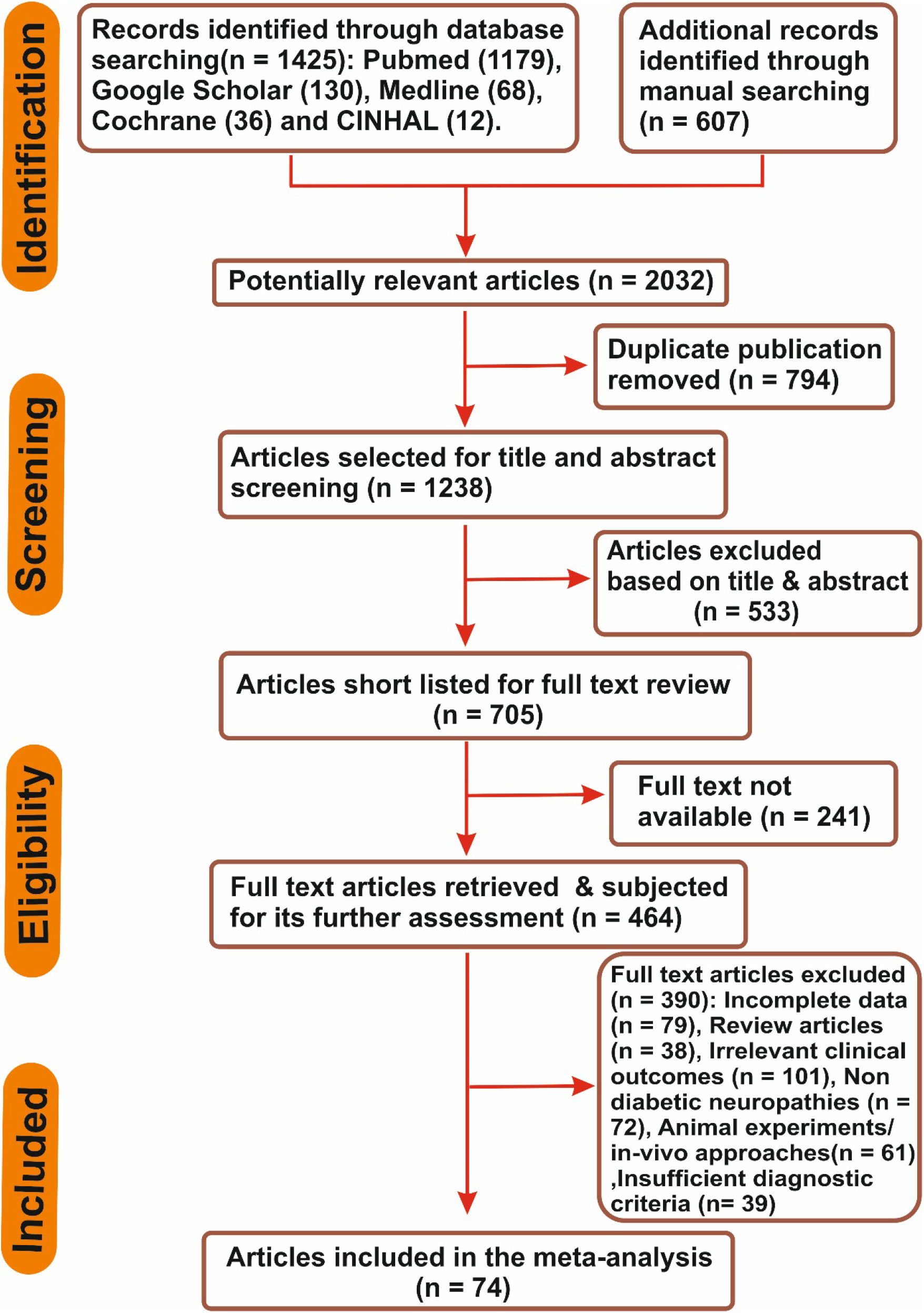
legend: PRISMA Flow Diagram Illustrating the Systematic Literature Search and Study Selection Process. A four-phase selection framework Identification, Screening, Eligibility, and Inclusion was employed following standard systematic review methodology. An initial database search across five electronic repositories (PubMed, n = 1,179; Google Scholar, n = 130; Medline, n = 68; Cochrane Library, n = 36; CINHAL, n = 12) yielded 1,425 records, supplemented by 607 manually identified articles, totalling 2,032 potentially relevant studies. Following removal of 794 duplicate publications, 1,238 articles underwent title and abstract screening, of which 533 were excluded for non-alignment with the predefined DPN pathophysiological framework. Of the 705 articles shortlisted for full-text review, 241 were inaccessible due to unavailability of full text, leaving 464 articles subjected to detailed eligibility assessment. A further 390 articles were excluded on the following grounds: irrelevant clinical outcomes (n = 101), non-diabetic neuropathies (n = 72), incomplete data (n = 79), animal or in vitro experimental models (n = 61), review articles or editorials (n = 38), and insufficient diagnostic criteria (n = 39). Ultimately, 74 studies satisfied all predefined inclusion criteria and were retained for quantitative meta-analysis.

The 74 included studies collectively enrolled 83,560 participants, of whom 23,998 were confirmed DPN patients. Sample sizes ranged from 10 to 37,375 participants, originating from 24 countries across six continents: Europe (n=37), Asia (n=23), North America (n=10), Latin America (n=1), Australia (n=1), and Africa (n=2). Diagnostic modalities comprised: Nerve Conduction Studies (NCS, n=31), Michigan Neuropathy Screening Instrument (MNSI, n=20), standard clinical neurological assessment (n=27), and nerve/skin biopsy (n=11). NOS quality assessment yielded: 5 studies scoring 9, 3 scoring 8, 6 scoring 7, 21 scoring 6, 19 scoring 5, 17 scoring 4, and 4 scoring 3 (Supplementary Table S2).

### Global Pooled Prevalence and Continent-Specific Subgroup Analysis

The global pooled prevalence of DPN was 57.42% (95% CI: 48.56–66.05%) under a random-effects model with Freeman–Tukey double arcsine transformation, with extreme between-study heterogeneity (I² = 99%, p < 0.001) (Fig. 2). Continent-specific subgroup meta-analysis revealed marked geographic variation (Q = 5.90, df = 2, p = 0.0524) (Table-1) (Supplementary Fig. S1). The pooled prevalence in Europe (36 studies) was 59.95% (95% CI: 45.81–73.33%; I² = 99.6%), substantially exceeding previously reported European estimates of 35% across five major European nations, 18.87% in Greece, 23.3% in Denmark, and 28% in the EURODIAB IDDM Complications Study [22–25]. In the Americas (11 studies), the pooled prevalence was 74.52% (95% CI: 46.27–94.66%; I² = 99.8%), markedly surpassing reported estimates of 28% for U.S. diabetic adults, 39.2% in U.S. cross-sectional cohorts, 14.6% in Mexico City, and 46.5% in Latin America and the Caribbean [26–29]. In Asia (23 studies), the pooled prevalence was comparatively lower at 45.44% (95% CI: 34.46–56.64%; I² = 98.9%), consistent with reported ranges of 28–37% in Japan, 67.7% in China, and 9.6–78% in India [30–32]. The pervasive extreme heterogeneity (I² > 98% throughout) reflects the influence of diverse population ethnicities, heterogeneous diagnostic criteria, variable case definitions, comorbidity burdens, and differential healthcare infrastructure across settings.

**Figure 2.**
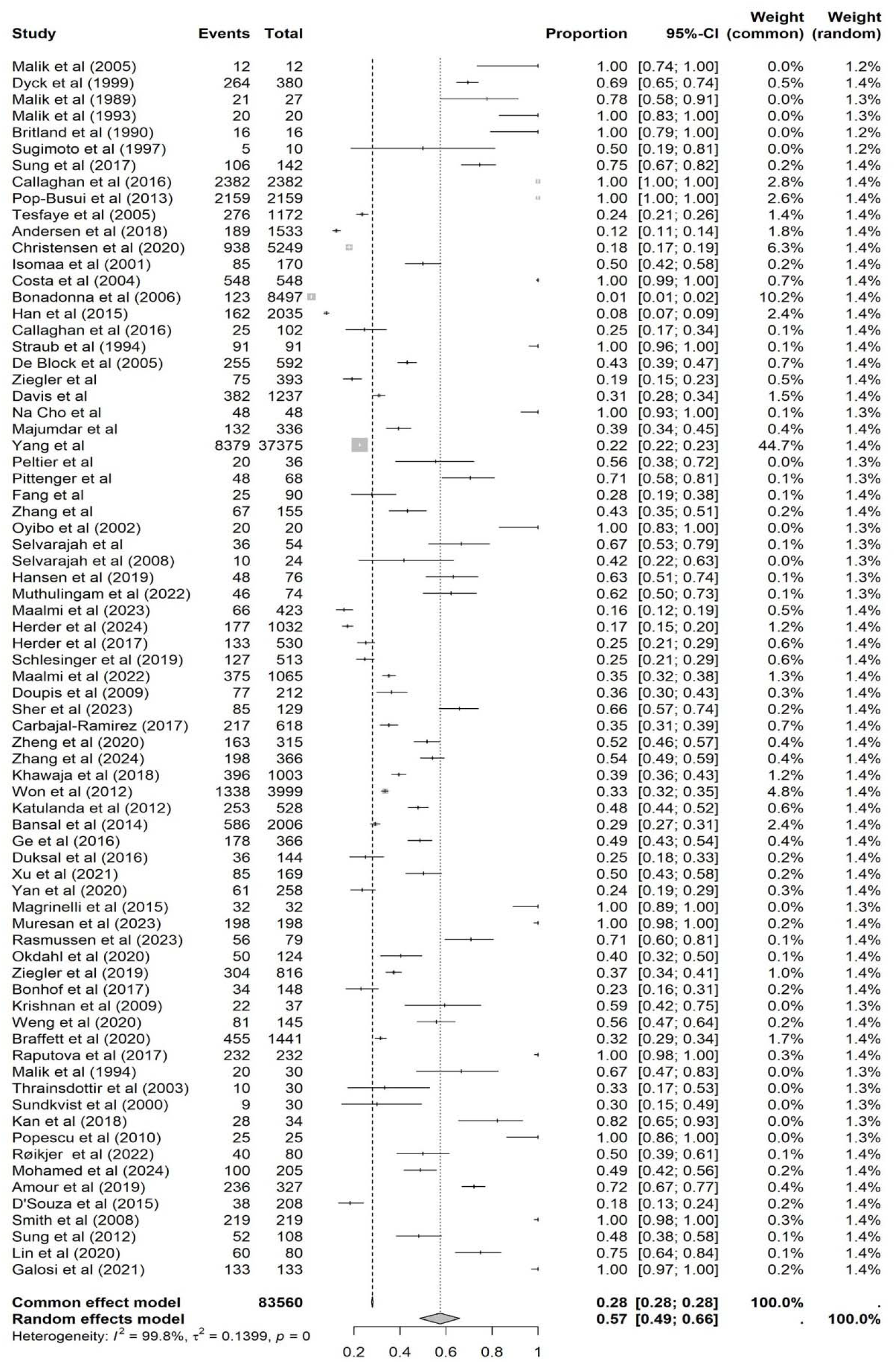
legend: Forest Plot Depicting the Global Pooled Prevalence of Diabetic Peripheral Neuropathy (DPN) Across 74 Studies. Each horizontal line represents an individual study, with the central square indicating the point prevalence estimate and the line width denoting the 95% confidence interval (CI). Square size is proportional to the assigned study weight under the random-effects model. The vertical dashed line represents the pooled prevalence estimate. The pooled results are presented at the bottom for both the common-effect model (proportion: 0.28; 95% CI: 0.28–0.28) and the random-effects model (proportion: 0.57; 95% CI: 0.49–0.66), with the latter considered the primary estimate given the substantial between-study heterogeneity. Heterogeneity was extreme across all included studies (I² = 99.8%, τ² = 0.1399, p = 0), necessitating the application of the random-effects model with Freeman–Tukey double arcsine transformation for variance stabilization prior to prevalence pooling. Study-level weights under the common-effect and random-effects models are displayed in the rightmost columns. The overall analysis incorporated 83,560 participants spanning 74 studies from 24 countries across six continents, with individual study proportions ranging from 0.01 (Bonadonna et al., 2006) to 1.00 across multiple studies, reflecting the substantial heterogeneity in DPN diagnostic criteria, study populations, and healthcare settings.

**Table 1.** Diabetic neuropathy associated continent based sub-group analysis of prevalence’s.

| Continent | Number of Studies | <u>Heterogeneity</u> |  |  | Q value for difference between sub-groups | p- value for difference between sub-groups |
| --- | --- | --- | --- | --- | --- | --- |
|  |  | Prevalence (95 %CI), % | tau^2 | tau |  |  |
| Europe | 37 | 59.95<br>(45.81 – 73.33) | 0.1710 | 0.4136 | 5.90 | 0.0524 |
| America | 11 | 74.52<br>(46.27 – 0.9466) | 0.1875 | 0.4331 |  |  |
| Asia | 23 | 45.44<br>(34.46 – 56.64) | 0.0629 | 0.2509 |  |  |

### Hallmark-Based Subgroup Analysis

#### Neuronal Damage

Neuronal damage subgroup analysis (26 studies; 2,835 participants; 1,279 DPN patients) encompassed axonal dysfunction, segmental demyelination, nerve fiber density reduction, sensory neuronal degeneration, impaired nerve regeneration, and neurotransmitter imbalance. Pooled prevalence trended upward with increasing hallmark burden: 0.54 (95% CI: 0.43–0.66) at score 0, 0.55 (95% CI: 0.39–0.71) at score 1, and 0.68 (95% CI: 0.44–0.88) at score ≥2, without achieving statistical significance (Q = 1.28, df = 2, p = 0.527) (Table-2). The absence of significant subgroup differentiation likely reflects incomplete electrophysiological characterization: 59.45% of studies did not employ NCS-based peripheral neurophysiological testing the diagnostic gold standard for DPN precluding rigorous structural nerve damage quantification. Additionally, currently approved pharmacological agents are fundamentally incapable of reversing established structural nerve injury, which may further obscure the relationship between neuronal damage severity and observed prevalence [3,5].

**Table 2.** neuropathy associated hallmark based sub-group analysis of prevalence’s.

| DPN<br>Hallmark | Sub-Group | Number<br>of<br>Studies | Heterogeneity |  |  | Q value<br>for<br>difference<br>between<br>sub-groups | p- value<br>for<br>difference<br>between<br>sub-groups |
| --- | --- | --- | --- | --- | --- | --- | --- |
|  |  |  | Prevalence<br>(95 %CI), % | tau^2 | tau |  |  |
| Neuronal<br>Damage | Sub-Group =<br>0 | 48 | 54.94<br>(43.21 – 66.4) | 0.1597 | 0.3997 | 1.28 | 0.5268 |
|  | Sub-Group =<br>1 | 13 | 55.69<br>(39.75 – 71.08) | 0.0615 | 0.2481 |  |  |
|  | Sub-Group =<br>≥2 | 13 | 68.51<br>(44.72 - 88.25) | 0.1453 | 0.3812 |  |  |
| Metabolic<br>Dysregulation | Sub-Group =<br>0 | 12 | 74.31<br>(53.14 – 91.02) | 0.1058 | 0.3252 | 3.95 | 0.1386 |
|  | Sub-Group =<br>1 | 21 | 59.55<br>(44.90 – 73.41) | 0.0976 | 0.3124 |  |  |
|  | Sub-Group =<br>2 | 41 | 51.43<br>(38.61 – 64.16) | 0.1643 | 0.4054 |  |  |
| Inflammation | Sub-Group =<br>0 | 55 | 59.13<br>(48.53 – 69.33) | 0.1475 | 0.3841 | 0.46 | 0.4953 |
|  | Sub-Group =<br>1 | 19 | 52.48<br>(35.48 – 69.20) | 0.1212 | 0.3482 |  |  |
| Micro-<br>vascular<br>Alteration | Sub-Group =<br>0 | 64 | 53.59<br>(44.20 – 62.85) | 0.1371 | 0.3703 | 15.78 | 0.0004 |
|  | Sub-Group =<br>1 | 6 | 72.58<br>(33.78 – 98.32) | 0.1275 | 0.3571 |  |  |
|  | Sub- | 4 | 92.31 | 0.0296 | 0.1719 |  |  |
|  | Group =<br>≥2 |  | (65.34 – 1.000) |  |  |  |  |

#### Metabolic Dysregulation

Metabolic dysregulation subgroup analysis (62 studies; 82,708 participants; 23,510 DPN patients) demonstrated a counterintuitive, inverse gradient: pooled prevalence decreased from 0.74 (95% CI: 0.53–0.91) at score 0, to 0.59 (95% CI: 0.44–0.73) at score 1, and 0.51 (95% CI: 0.38–0.64) at score 2, without statistical significance (Q = 3.95, df = 2, p = 0.139) (Table-2). This paradoxical finding is mechanistically interpretable. Study operationalization was restricted to two features (hyperglycemia and dyslipidemia), failing to capture the contemporary multifactorial metabolic landscape of DPN which encompasses insulin resistance, mitochondrial dysfunction, oxidative stress, hypertension, and central obesity [5,33]. Three converging lines of evidence support this interpretation. First, patients with adequate glycemic control (HbA1c < 5.4%) remain susceptible to DPN progression, underscoring the therapeutic inadequacy of glycemia-centric management [3,5,10]. Second, glycemic variability a dimension of metabolic dysregulation independent of mean glucose lacks both a gold-standard assessment tool and effective pharmacological targeting [34]. Third, metabolic syndrome (MetS), defined by the co-clustering of central obesity, insulin resistance, dyslipidemia, hypertriglyceridemia, and hypertension, represents a more mechanistically potent activator than isolated hyperglycemia or dyslipidemia [3,5,33]. Consistent with this interpretation, our meta-analysis identified MetS as a near-significant DPN risk factor (OR: 1.21; 95% CI: 0.99–1.49; p = 0.055), while FPG (OR: 1.77; 95% CI: 0.90–3.49; p = 0.088) and dyslipidemia (OR: 1.13; 95% CI: 0.57–2.25; p = 0.648) were individually non-significant (Table-3).

**Table 3.** Pooled risk factors linked with diabetic peripheral neuropathy development and progression.

| Risk Factors | No. of study involved | Combined OR (95% CI) | I<sup>2</sup> | τ<sup>2</sup> | p- value |
| --- | --- | --- | --- | --- | --- |
| Foot Ulcers | 3 | 5.81 (1.59 – 21.24) | 0.0 | 0.000 | 0.0280 |
| Peripheral Vascular Disease | 4 | 3.70 (2.22 – 6.16) | 0.0 | 0.007 | 0.00384 |
| Diabetic Retinopathy | 9 | 2.37 (1.77 – 3.17) | 0.8 | 0.109 | 0.000138 |
| Insulin Treatment | 4 | 1.82 (0.71 – 4.66) | 0.8 | 0.245 | 0.134 |
| FPG | 9 | 1.77 (0.90 – 3.49) | 0.9 | 0.637 | 0.0878 |
| Hypertension | 9 | 1.71 (1.29 – 2.26) | 0.6 | 0.051 | 0.00231 |
| BMI | 10 | 1.63 (0.91 – 2.93) | 0.9 | 0.522 | 0.0927 |
| Diabetes Duration | 13 | 1.59 (1.07 – 2.37) | 0.9 | 0.361 | 0.0262 |
| Diabetic Nephropathy | 4 | 1.55 (0.73 – 3.31) | 0.5 | 0.105 | 0.163 |
| Cardiovascular Disease | 8 | 1.54 (1.31 – 1.80) | 0.5 | 0.021 | 0.000344 |
| Male Population | 8 | 1.52 (1.19 – 1.94) | 0.4 | 0.033 | 0.00526 |
| Female Population | 8 | 1.47 (0.52 – 4.15) | 0.8 | 0.566 | 0.365 |
| Patient Age | 12 | 1.43 (1.07 – 1.91) | 0.9 | 0.126 | 0.0200 |
| HbA1c | 11 | 1.40 (0.86 – 2.29) | 0.9 | 0.409 | 0.151 |
| History of Smoking | 9 | 1.38 (0.82 – 2.34) | 0.7 | 0.266 | 0.192 |
| Insulin Resistance | 2 | 1.31 (0.17 – 10.09) | 0.4 | 0.03 | 0.339 |
| Height | 5 | 1.26 (1.00 – 1.57) | 0.9 | 0.023 | 0.0481 |
| Waist Circumference | 9 | 1.26 (1.04 – 1.54) | 0.7 | 0.038 | 0.0249 |
| Metabolic Syndrome | 3 | 1.21 (0.99 – 1.49) | 0.0 | 0.000 | 0.0552 |
| Triglycerides | 9 | 1.15 (0.97 – 1.36) | 0.7 | 0.017 | 0.104 |
| Dyslipidemia | 5 | 1.13 (0.57 – 2.25) | 0.7 | 0.186 | 0.648 |
| DBP | 2 | 1.09 (0.31 – 3.80) | 0.9 | 0.018 | 0.536 |
| Alcohol Consumption | 6 | 1.08 (0.94 – 1.25) | 0.5 | 0.003 | 0.195 |
| Total Cholesterol | 5 | 1.03 (0.94 – 1.13) | 0.3 | 0.001 | 0.376 |
| SBP | 5 | 1.01 (0.90 – 1.14) | 0.9 | 0.008 | 0.766 |
| LDL | 2 | 1.01 (0.59 – 1.73) | 0.1 | 0.002 | 0.863 |
| HDL | 6 | 0.89 (0.79 – 1.00) | 0.5 | 0.000 | 0.0457 |
| Physical Activity | 2 | 0.57 (0.00 – 93.88) | 0.5 | 0.173 | 0.394 |

#### Neuroinflammation

Neuroinflammation subgroup analysis (19 studies; 10,682 participants; 2,892 DPN patients) demonstrated a modest, non-significant decline in pooled prevalence from 0.59 (95% CI: 0.48–0.69) at score 0 to 0.52 (95% CI: 0.35–0.69) at score 1 (Q = 0.46, df = 1, p = 0.495) (Table-2). This finding is plausibly attributable to divergent neuroinflammatory profiles across diabetes types: in type 2 diabetes DPN, neuroinflammatory biomarker levels are paradoxically diminished relative to controls independent of neuropathic pain status as demonstrated across 71 serum biomarkers [35], whereas type 1 diabetes DPN is typically associated with elevated circulating neuroinflammatory biomarkers [36]. Since 17 of 19 included studies involved type 2 diabetes cohorts, their attenuated neuroinflammatory signatures may substantially underestimate the true neuroinflammatory burden at the global DPN level. Furthermore, predominant cross-sectional study designs preclude causal inference regarding the temporal relationship between subclinical inflammation and neuropathy development, and limited sample sizes reduce statistical power for phenotypic differentiation [37].

#### Microvascular Alteration

Microvascular alteration subgroup analysis (10 studies; 786 participants; 493 DPN patients) demonstrated a striking, monotonic escalation in pooled DPN prevalence with increasing hallmark severity: 0.53 (95% CI: 0.44–0.62) at score 0, 0.72 (95% CI: 0.33–0.98) at score 1, and 0.92 (95% CI: 0.65–1.00) at score ≥2 achieving statistical significance under random-effects subgroup testing (Q = 15.78, df = 2, p = 0.0004) (Table-2). This dose-response gradient represents the sole statistically significant hallmark subgroup result, providing robust meta-analytic evidence for the mechanistic primacy of endoneurial vascular pathology encompassing capillary basement membrane widening, pericyte loss, and endothelial dysfunction in determining clinical DPN burden [2,3,18,38]. These findings align with established mechanistic evidence implicating endoneurial ischemia-hypoxia, capillary rarefaction, and impaired nerve perfusion in perpetuating axonal energy deprivation and structural nerve injury [38–40]. From a therapeutic standpoint, these data argue compellingly for prioritized pharmacological targeting of microvascular mechanisms, including nerve blood flow restoration, neuroprotection against ischemia-hypoxia, and capillary density improvement potentially achievable through vasodilatory agents such as alpha-adrenergic receptor blockers, prostaglandin E1 analogues, endothelin receptor antagonists, angiotensin II receptor blockers, and angiotensin-converting enzyme inhibitors [38,40].

#### Risk Factor Assessment

Across 74 studies encompassing 83,560 participants, 27 clinical and demographic risk factors were systematically analyzed. Random-effects meta-analysis identified 12 statistically significant risk factors (p ≤ 0.05) (Table-3). The four highest-significance risk factors were diabetic comorbidities: diabetic retinopathy (OR: 2.37; 95% CI: 1.77–3.17; p = 0.000138), cardiovascular disease (OR: 1.54; 95% CI: 1.31–1.80; p = 0.000344), hypertension (OR: 1.71; 95% CI: 1.29–2.26; p = 0.00231), and peripheral vascular disease (PVD) (OR: 3.70; 95% CI: 2.22–6.16; p = 0.00384). Notably, PVD is independently reported here as a significant DPN risk factor for the first time in a meta-analytic framework; its mechanistic basis involves limb ischemia-driven vasa nervorum compromise synergizing with DPN-associated sensorimotor deficits to exponentially amplify risk for DFU and subsequent amputation [41,42]. Foot ulcers (OR: 1.71; 95% CI: 1.29–2.26; p = 0.00231) represent an additional clinically consequential bidirectional complication-DPN association.

Among patient-intrinsic factors, diabetes duration (OR: 1.59; 95% CI: 1.07–2.37; p = 0.026) and patient age (OR: 1.43; 95% CI: 1.07–1.91; p = 0.020) confirmed their established pathogenic roles, with prolonged hyperglycemic exposure in older individuals preferentially activating the neuronal damage hallmark through electrophysiological deterioration, reduced nerve conduction velocities, and structural axonopathy [43,44]. Two risk factors are newly confirmed as statistically significant in this meta-analysis: male sex (OR: 1.52; 95% CI: 1.19– 1.94; p = 0.005) and height (OR: 1.26; 95% CI: 1.00–1.57; p = 0.048) both previously non-significant in earlier meta-analyses [14]. The biological plausibility of height as a DPN risk factor rests on the greater metabolic vulnerability of longer peripheral axons to ischemia-hypoxia and metabolic-toxic injury, with axonal length independently predicting DPN severity through both neuronal damage and microvascular alteration hallmarks [47,48]. Male sex predisposition likely reflects a convergence of sex-specific metabolic risk profiles, higher prevalence of electrophysiological abnormalities, elevated microvascular alteration hallmark activation, and greater burden of distal symmetric polyneuropathy (DSPN) [49,50].

Fifteen risk factors were non-significant: FPG, HbA1c, BMI, SBP, DBP, triglycerides, total cholesterol, LDL, dyslipidemia, insulin treatment, nephropathy, female sex, alcohol consumption, physical activity, and smoking (Table 3). Divergences from Y. Tao et al. (2025) and T.M. Fakkel et al. (2020) for HbA1c, BMI, SBP, female sex, nephropathy, and smoking [11,14] likely reflect differences in study composition and the substantially broader geographic and methodological diversity of the present dataset.

#### Publication Bias and Sensitivity Analysis

Publication bias was assessed using contour-enhanced funnel plots and Egger’s regression tests (Supplementary Fig. S2). Eight risk factors (FPG, BMI, HDL, triglycerides, height, waist circumference, insulin treatment, alcohol consumption) exhibited greater funnel asymmetry, raising concern for potential small study effects. Ten risk factors (HbA1c, diabetes duration, SBP, hypertension, CVD, dyslipidemia, diabetic retinopathy, nephropathy, male sex, smoking) demonstrated asymmetry patterns, with moderate concern for small study effects. PVD demonstrated the lowest publication bias risk with the most near-symmetric funnel distribution, supporting the reliability of its pooled effect estimate. For diabetes duration, notable funnel asymmetry confirmed by significant Egger’s test warrants cautious interpretation. HbA1c and BMI showed relatively symmetric funnels with non-significant Egger’s tests, providing no strong statistical evidence of small study-effects, though the possibility of undetected bias cannot be excluded. For covariates with sparse contributing studies (female sex, patient age, total cholesterol, LDL, DBP, physical activity), apparent asymmetry most likely reflects sampling variability rather than systematic bias.

## DISCUSSION

The onset and progression of DPN impose a substantial multi-dimensional disease burden encompassing physical (neuropathic pain, sensory deficit, falls, fractures), complication-driven (DFU, non-traumatic amputation, infection), and psychosocial (depression, anxiety, reduced health-related quality of life) dimensions [1–6]. Prior meta-analyses have primarily focused on DPN prevalence estimation and risk factor identification [11–15], with a conspicuous absence of mechanistic meta-analytic investigation a void that has critically impeded timely diagnosis, avoidance of under-diagnosis, mitigation of treatment adverse effects, and optimization of treatment response [5–10]. Existing conventional therapies are largely incapable of reversing established nerve injury, effectively controlling neuropathic pain, or providing disease-modifying benefit [10,51,52].

The present systematic review and meta-analysis comprehensively addresses these gaps. Based on 74 studies comprising 83,560 participants, it constitutes the largest and most mechanistically detailed DPN meta-analysis to date, and the first to quantitatively validate a hallmark-based mechanistic framework for DPN pathogenesis. The global pooled DPN prevalence of 57.42% indicating that more than one in two individuals across included studies manifest DPN substantially exceeds previously published estimates of 46.7% (Y. Tao et al., 2025; 41 studies, 36,983 participants) and 30% (J. Sun et al., 2020; 29 studies, 50,112 participants) [11–15]. This higher estimate reflects the broader geographic sampling across six continents and 24 countries, explicitly incorporating low-resource settings (Sri Lanka, Tanzania, Egypt, Bosnia and Herzegovina, Romania) where DPN is systematically under-recognized and under-treated.

The microvascular alteration hallmark emerged as the sole statistically significant mechanistic subgroup determinant of DPN prevalence (Q = 15.78, p = 0.0004), with pooled prevalence escalating from 53% to 92% across increasing microvascular severity scores. This dose-response gradient constitutes novel meta-analytic evidence for the mechanistic primacy of endoneurial vascular pathology in clinical DPN burden, and directly supports microvascular-targeted pharmacotherapy as a priority approach. Conversely, the absence of significant subgroup differentiation for neuronal damage, metabolic dysregulation, and neuroinflammation hallmarks does not negate their pathophysiological relevance; rather, it reflects methodological heterogeneity and data gaps inherent to available clinical datasets — including incomplete electrophysiological phenotyping, cross-sectional designs precluding causal inference, and narrow operationalization of complex multi-feature hallmarks.

The risk factor meta-analysis identified 12 significant DPN determinants, including two novel meta-analytic findings: PVD as the strongest individual risk factor (OR: 3.70) and height as a previously under-recognized contributor (OR: 1.26). The identification of PVD as the highest-effect-size risk factor mechanistically links systemic arteriolar disease with endoneurial ischemia a direct activator of the microvascular alteration hallmark and has immediate clinical implications for integrated vascular-neurological DPN risk stratification. The confirmation of male sex as a significant risk factor substantiates sex-specific neuropathy vulnerability, potentially shaped by androgen-related metabolic and microvascular risk interactions [49,50]. The non-significance of HbA1c, BMI, SBP, and nephropathy contrasting with prior meta-analyses [11,14] may reflect the heterogeneous clinical profiles of the expanded, geographically diverse study cohort, and underscores the inadequacy of single-metric glycemic or metabolic targets as universal DPN risk predictors. These discordant findings collectively reinforce the concept that DPN is a mechanistically heterogeneous, multifactorial disease requiring hallmark-stratified, patient-specific risk assessment rather than population-averaged risk benchmarks.

### Study Limitations

Several limitations warrant careful consideration. First, the predominance of cross-sectional study designs precludes causal inference and limits evaluation of longitudinal risk factor effects. Second, 59.5% of studies did not employ NCS-based peripheral neurophysiological testing, potentially leading to under-ascertainment of peripheral nerve damage. Third, 54 studies lacked standardized MNSI-based neuropathic symptom assessment, reducing cross-study comparability. Fourth, substantial sample size variability (10–37,375 participants) introduces heterogeneity in effect estimate precision. Fifth, pain-symptom data were largely absent across included studies, precluding meta-analytic assessment of PDPN. Sixth, inclusion of type 1, type 2, and mixed-diabetes cohorts may introduce selection bias in hallmark-prevalence associations. Seventh, several key risk factors (DBP, MetS, foot ulcers) were reported in three or fewer studies, constraining statistical reliability of their pooled ORs.

### Study Strengths

Our meta-analysis offers several methodologically and scientifically distinctive strengths. It is the first to propose and quantitatively validate a hallmark-based mechanistic framework for DPN pathogenesis encompassing neuronal damage, microvascular alteration, metabolic dysregulation, and neuroinflammation using clinical data from 74 studies. Geographically, it achieves the broadest coverage to date (six continents, 24 countries), including low-income and under-resourced nations rarely represented in prior DPN meta-analyses. The composite enrollment of 83,560 participants provides sufficient statistical depth for robust pooled estimation and subgroup analysis. Diagnostic diversity across NCS, MNSI, clinical neurological assessment, and nerve/skin biopsy with 51 studies employing dual-method ascertainment enhances clinical validity and generalizability. Additionally, incorporation of MRI-based brain volumetric data reflects CNS contributions to DPN pathogenesis, and specific inclusion of small fiber neuropathy studies capturing PDPN and early-stage DFU risk extends beyond what conventional NCS and MNSI can detect.

### Future Implications and Conclusion

This meta-analysis provides the most comprehensive quantitative synthesis to date of global DPN prevalence, hallmark-based mechanistic stratification, and multi-covariate risk factor profiling. Given the inherent heterogeneity and multifactorial etiology of DPN, the precision medicine paradigm grounded in hallmark-stratified phenotyping and mechanism-targeted pharmacotherapy represents the most scientifically justified future direction [1]. Our hallmark-based subgroup framework provides a methodological foundation for systematic phenotype characterization in DPN, enabling the dissection of neuronal damage, microvascular, metabolic, and neuroinflammatory disease endotypes both individually and in combination. Future research should prioritize: (i) development of large-scale, prospective, multicenter DPN cohorts stratified by hallmark severity scores; (ii) discovery and external validation of hallmark-specific biomarkers (imaging, electrophysiological, molecular) for early phenotypic diagnosis; (iii) design of mechanism-targeted clinical trials enrolling patients stratified by dominant hallmark burden; and (iv) integration of multi-omics data to identify patient-specific molecular drivers across hallmark categories. The 12 statistically significant risk factors identified particularly the novel meta-analytic confirmation of PVD (OR: 3.70), MetS (OR: 1.21), male sex, and height provide actionable targets for precision lifestyle and pharmacological intervention. A patient-centric, comorbidity-aware assessment of these risk factors, linked to individualized hallmark-based disease mechanisms, holds potential to realize lifestyle precision medicine for DPN and to identify disease-modifying treatment strategies capable of altering the natural history of DPN progression.

In conclusion, this systematic review and meta-analysis documents a global pooled DPN prevalence of 57.42%, substantially exceeding prior estimates, with important continent-specific gradients. Microvascular alteration emerges as the dominant, statistically validated mechanistic hallmark of DPN pathogenesis, with a striking dose-response relationship between hallmark severity and DPN prevalence. Twelve significant risk factors are identified, including the novel meta-analytic confirmation of PVD, MetS, male sex, and height as DPN determinants. Although three of four hallmarks did not produce statistically significant subgroup differences attributed to methodological heterogeneity, data limitations, and hallmark operationalization constraints the overall mechanistic framework demonstrates clinical and biological coherence. Substantial future work is required through large-scale, multicenter, prospective study designs incorporating standardized hallmark-aligned phenotyping, external validation of identified biomarkers, and rigorous evaluation of multi-modal, hallmark-targeted management strategies.

## Conflict of interest statement

The authors declare no potential conflict of interest at any level.

## Data availability statement

Current study utilizes publicly accessible data from published studies; therefore no original data are present for sharing.

## Funding

This research did not receive any specific grant from funding agencies in the public, commercial, or not-for-profit sectors.

## Author contributions

DD, DS, SPH and BG designed the study. DD reviewed the literature. DD, DS, SPH, RG and BG interpreted the data. RG and BG performed statistical analysis. DD, DS, AB and SPH provided medical insights and epidemiological analysis. DD and DS prepared the figures. DD, DS, RG and SPH wrote the first draft of the manuscript. All authors contributed in data interpretation, validation, manuscript revision and approval of final manuscript. SPH supervised the overall study.

## Supporting information

Suupplemental Figure S1

Suupplemental Figure S2

Suupplemental Table S3

## Data Availability

All data produced in the present work are contained in the manuscript

## Acknowledgements

The author thanks Dr. Partha Chakrabarty from CSIR-IICB, Kolkata for his critical support in conceptualization, verification and manuscript evaluation.

---

**Supplementary Figure S1: Forest plot of the pooled prevalence of peripheral diabetic neuropathy, stratified by continent.** Forest plot showing the prevalence of peripheral diabetic neuropathy (DN) reported in each of the included studies, grouped by continent (Europe, America, and Asia). For each study, the number of DN cases, the total sample size, and the corresponding effect estimate with 95% confidence interval (CI) are shown (inverse-variance method, random-effects model). Diamonds represent the pooled prevalence estimate for each continental subgroup and for the overall pooled analysis (95% CI). Heterogeneity within each subgroup is reported as Tau², Chi², and I² statistics, and the test for subgroup differences (Chi² test, df = 2) is shown below the overall estimate. The x-axis represents the prevalence of DN.

**Supplementary Figure S2: Contour-enhanced funnel plots evaluating publication bias for risk factors associated with peripheral diabetic neuropathy.** Contour-enhanced funnel plots (log odds ratio vs. standard error) assessing small-study effects and potential publication bias for each of the 28 risk factors evaluated in the meta-analysis: (A) alcohol consumption, (B) any retinopathy, (C) BMI, (D) cardiovascular disease, (E) diastolic blood pressure, (F) diabetes duration, (G) dyslipidemia, (H) female sex, (I) fasting plasma glucose, (J) foot ulcers, (K) HbA1c, (L) HDL cholesterol, (M) height, (N) history of smoking, (O) hypertension, (P) insulin resistance, (Q) insulin treatment, (R) LDL cholesterol, (S) male sex, (T) metabolic syndrome, (U) nephropathy, (V) patient age, (W) peripheral vascular disease, (X) physical activity, (Y) systolic blood pressure, (Z) total cholesterol, (AA) triglycerides, and (AB) waist circumference. In each panel, individual studies are plotted as black dots; the solid vertical line indicates the pooled log odds ratio and the dashed vertical line indicates the null value (log OR = 0). Shaded contour regions denote conventional significance thresholds (p < 0.10, p < 0.05, p < 0.01), allowing visual distinction between asymmetry attributable to publication bias and asymmetry arising from genuine heterogeneity.

**Supplementary Table 1: Characteristics, quality assessment, and reference list of studies included in the systematic review and meta-analysis.** Sheet S1 lists the full bibliographic reference for each included study, identified by Study ID (first author and publication year). Sheet S2 summarizes the key characteristics of each study, including country of origin, total sample size, number of patients with peripheral diabetic neuropathy (DPN), study design, the DPN-related hallmark mechanism(s) assessed (e.g., neuronal damage, micro-vascular alterations, metabolic dysregulation, inflammation), and methodological quality as scored using the Newcastle–Ottawa Scale (NOS).

## Notes

### Competing Interest Statement

The authors have declared no competing interest.

