## Supplementary figures and images for "Prevalence, Hallmark-Based Mechanisms, and Risk Factors of Peripheral Diabetic Neuropathy: A Systematic Review and Meta-Analysis"

### Suupplemental Figure S1

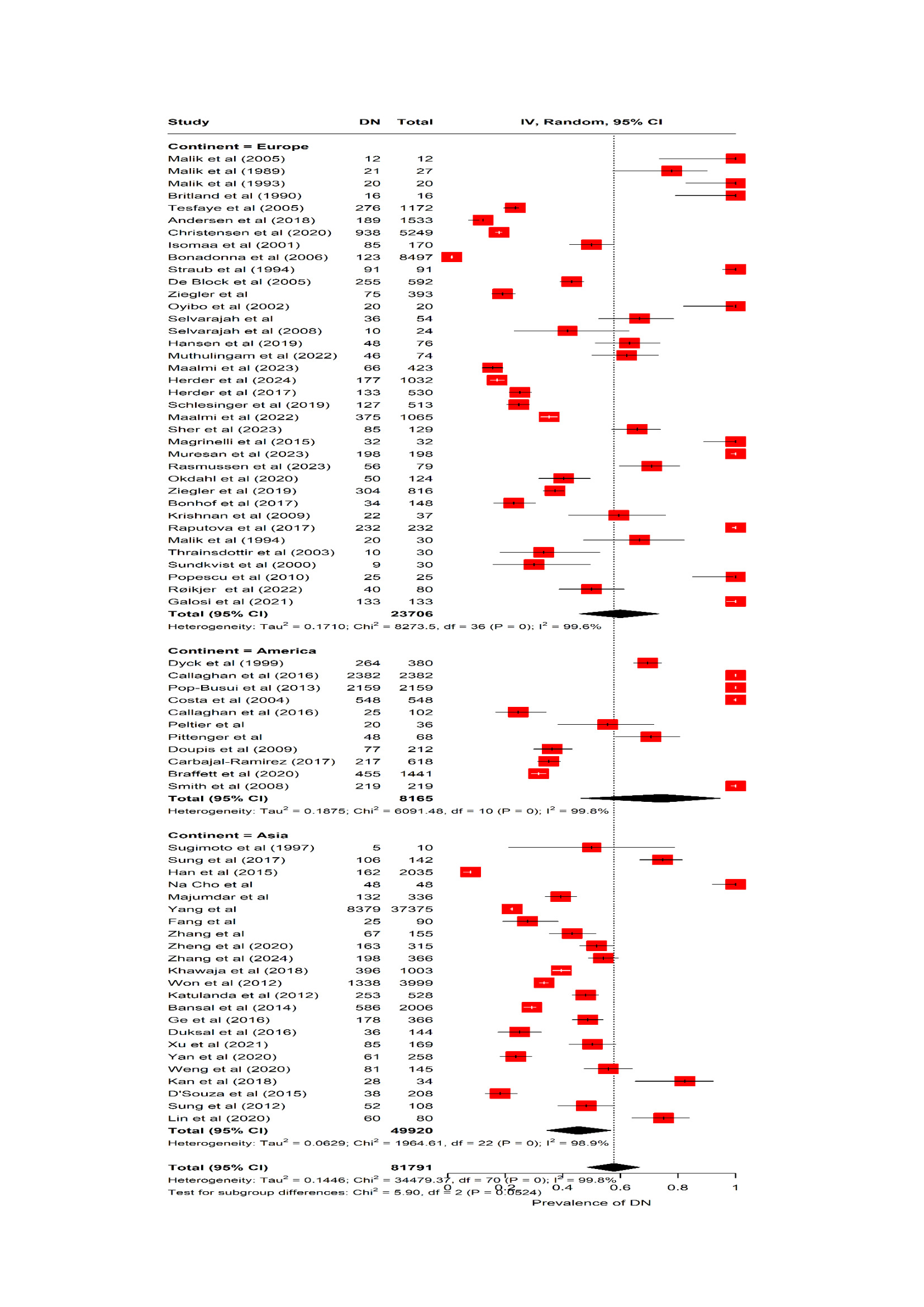

### Suupplemental Figure S2

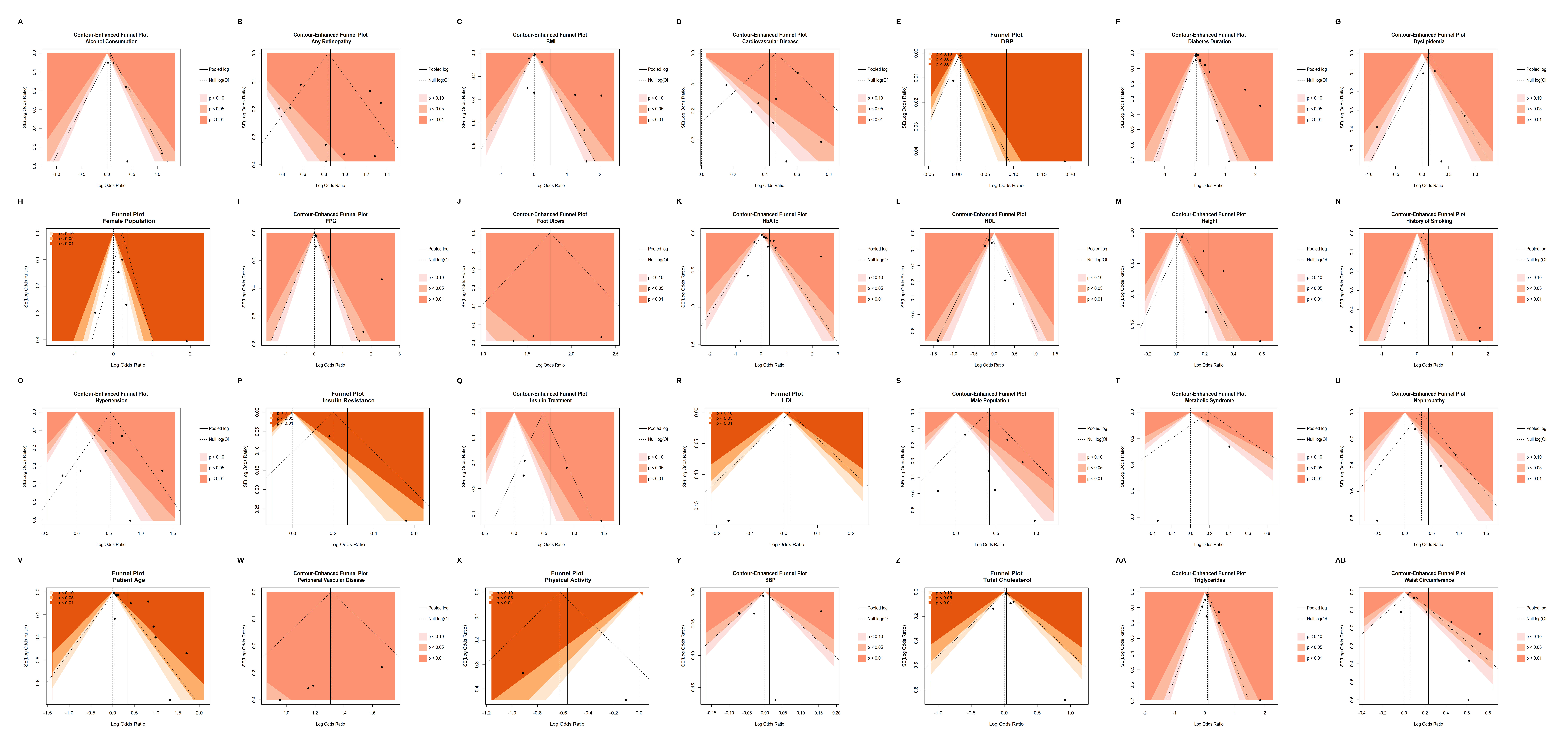
